# Screening the use of DCA for the treatment of cancer in adult patients: a scoping review protocol

**DOI:** 10.1101/2022.09.07.22279231

**Authors:** Cecilia Bianchi, Romina P Martinelli, Viviana R Rozados, O Graciela Scharovsky

## Abstract

Sodium dichloroacetate (DCA), a small-molecular-weight drug, is capable to shift the tumor metabolism by reactivating the oxidative phosphorylation in the mitochondria. As a consequence, apoptosis resistance in cancer cells decreased. Numerous studies have been performed in cell lines and animal models, to unveil its efficacy as an antitumor drug. It is also being studied in patients with different types of tumors, with a variety of doses and administration schedules. Even so, the lack of controlled studies makes the safety profile a concern. This scoping review aims to explore the existing scientific literature to provide an overview of the use of DCA (any dose, frequency, or route of administration) in adults with cancer. The search will be performed in MEDLINE/PubMed, and LILACS databases.

Additional data sources will be consulted. Two reviewers will screen all identified records for relevance, and chart the data using a data charting form. Findings will be reported according to PRISMA for Scoping Reviews (PRISMA-ScR). No quality assessment will be performed.

## 1 Rationale

In the last years, drug repurposing (DR), the strategy for identifying new uses for approved drugs, has become a strategy of great relevance due to the expensive and time/consuming traditional drug discovery processes. This strategy has been used in oncology for a long time. The first chemotherapy drugs, chlorambucil (Leukeran) and busulfan (Myleran) were developed from the ‘mustard gas’ used during the First and Second World Wars.

Straightaway, the knowledge of the chemical structure of drugs, as well as their mechanisms of action against different metabolic pathways, or on changes in gene expression, has allowed them to be assigned a new therapeutic purpose in different types of diseases for which they were not designed [1-4].

Moreover, the metabolism-oriented approach is a promising field for cancer therapy. Typically, tumor cells increased glucose uptake to generate lactate even in the presence of oxygen, a phenomenon known as the Warburg effect or aerobic glycolysis [5]. The pyruvate dehydrogenase kinase (PDK) is a fundamental enzyme controlling glycolysis and mitochondrial oxidative phosphorylation (OXPHOS) [6]. PDK is able to shut down the mitochondrial OXPHOS inhibiting pyruvate dehydrogenase (PDH), the enzyme catalyzes the oxidative conversion of pyruvate into acetyl coenzyme A in mitochondria [7]. Sodium dichloroacetate (DCA), widely used for the treatment of congenital lactic acidosis, is capable to shift the tumor metabolism by reactivating the oxidative phosphorylation in the mitochondria. As a consequence, apoptosis resistance in cancer cells decreased [8]. Based on this, the antitumor capacity of this compound has been studied with the aim of being used in the treatment of cancer [9-12]. Numerous studies have been performed in cell lines and animal models, to unveil its efficacy as an antitumor drug. It is also being studied in patients with different types of tumors, with a variety of doses and administration schedules. Even so, the lack of controlled studies makes the safety profile a concern.

For these reasons, this scoping review aims to explore the existing scientific literature to provide an overview of the use of DCA (any dose, frequency, or route of administration) in adults with cancer.

### 2 Objetives

Making allowance for the aim of this work, we set out the following question: What is known in the existing literature about the use of DCA for the treatment of cancer in adult patients?

The objectives of this scoping review are

i. To map the published literature reporting the use of DSA in cancer treatment
ii. To list the spectrum of tumors treated with DCA
iii. To indicate the dosis and frequency employed
iv. To summarize effectiveness and the main side effects reported during the follow-up

## 3 Methods

This scoping review protocol was developed following the methodological framework introduced by Arksey and O’Malley [13] and it was reported according to the PRISMA for Scoping Reviews (PRISMA-ScR) [14]

## 3.1 Stage 1: identifying the research question

In order to formulate the research question, the “PICO” methodology was used.

Population

Adults diagnosed with any type of cancer.

Intervention

We will include publications reporting dichloroacetate sodium as a cancer treatment in combination or not with other drugs.

Outcomes

The effectiveness and adverse effects will be taken into account

Types of studies

All types of study design will be included.

### 3.2 Stage 2: identifying relevant studies

The search will be conducted in MEDLINE/PubMed, and LILACS databases.

The search strategy will include Medical Subject Headings (MeSH) terms, and their relevant synonyms joined with Boolean operators. Searches will not be restricted by publication date, place, or type of study. EPISTEMONIKOS, and Cochrane Library databases will be explored to identify systematic reviews and meta-analyses as additional sources of primary studies. Search results will be downloaded in Comma-Separated Values (CSV) format on a single day.

The search strategy will be the following:

(Dichloroacetic Acid OR “dichloroacetate sodium” OR dichloroacetate OR “sodium dichloroacetate”) AND adult AND (cancer OR Neoplasms)

Additional studies will be added by scanning references of relevant articles.

#### 3.3 Stage 3: study selection

After removing duplicates, two researchers (CB and RPM) will screen the results at the title/ abstract level and second at the full-text level independently. Discrepancies will be resolved by consensus between the two reviewers. Articles written in English, Spanish, Portuguese, Italian, and French will be included.

### Inclusion criteria

All types of study design will be included.

Publications reporting on adult patients diagnosed with any type of cancer.

Publications reporting dichloroacetate sodium treatment in combination or not with other drugs.

### Exclusion criteria

Publications reporting on children or pregnant women. Studies on cell lines.

### 3.4 Stage 4: charting the data

We developed a data charting form with the following charting categories:

1. authors;
2. year of publication;
3. design;
4. Founding;
5. Age range;
6. Gender;
7. Disease;
8. DCA composition;
9. DCA dosis;
10. DCA route of administration;
11. Treatment duration;
12. Other drugs;
13. Follow up;
14. Adverse events;
15. Serious adverse events;
16. Outcome measures.

This form could be correct after being tested on 10 random studies. Data extraction will be independently performed by two researchers (CB and RPM), and discrepancies will be resolved by consensus.

### 3.5 Stage 5: collating, summarizing, and reporting the results

The PRISMA flowchart will be used to present the study selection process [15]. The data from each paper will be mapped in order to have a general idea of the use of DCA in the treatment of cancer.

Data will be presented in descriptive form. The quantitative results will be presented as appropriate in a chart or table form.

## Data Availability

All data produced in the present work are contained in the manuscript

## 4 Ethics and dissemination

The scoping review does not require ethical approval. The findings will be submitted for publication in scientific journals.

## 5 Contributions of authors

RPM drafted the protocol. RPM and CB developed the search strategy and conducted electronic searches. VRR and OGS contributed with content expertise and helped to develop the protocol.

